# Six-month mortality has decreased for patients with curative treatment for head and neck cancer in Sweden

**DOI:** 10.1101/2023.12.16.23300079

**Authors:** Charbél Talani, Anders Högmo, Göran Laurell, Antti Mäkitie, Lovisa Farnebo

## Abstract

**Background:** In general, survival outcome for head and neck cancer (HNC) patients has improved. However, mortality within six months after diagnosis with curative intent remains high. The aim of this study was to identify risk factors for early death among patients with curative treatment, and furthermore, to analyze whether the risk of early death changed over the recent years.

**Material and Method:** This real-world, population-based, nationwide study from the Swedish Head and Neck Cancer Register (SweHNCR) included 16,786 patients with curative treatment decision at the multidisciplinary tumor board from 2008 to 2020.

**Results:** During the study period a total of 618 (3.7%) patients with curative-intended treatment died within six months from diagnosis. Patients diagnosed between 2008–2012 had a six-month mortality rate of 4.7% compared to 2.5% for patients diagnosed 2017–2020, indicating a risk reduction of 53% (p <0.001) for death within six months. The mean time to radiation therapy in the 2008–2012 cohort was 38 days, compared to 22 days for the 2017– 2020 cohort, (p<0.001). The mean time to surgery from diagnosis was 22 days in 2008–2012, compared to 15 days for the 2017–2020 cohort, (p <0.001).

Females had a 20% lower risk of dying within six months compared to males (p =0.013). For every year older the patient was at diagnosis, a 4.8% (p <0.001) higher risk of dying within six months was observed. Patients with a WHO score of 1 had approximately 2.4-times greater risk of early death compared to WHO 0 patients (p <0.001). The risk of early death among WHO 4 patients was almost 28 times higher than for WHO 0 patients (p <0.001)

**Conclusions:** We found that the risk of early death decreased significantly from 2008 to 2020. During this period the mean time to the start of treatment, was significantly reduced both for surgery and oncological treatment regimes. Among patients with a curative treatment intention, increased risk of early death was associated with male sex, older age, advanced disease, increased WHO score, and a hypopharyngeal tumor site.

**Brief description:** *The risk of early death decreased significantly from 2008 to 2020.

*Time from diagnosis to treatment decreased between 2008 to 2020.

*Among patients with a curative treatment intention, increased risk of early death was associated with male sex, older age, advanced disease, increased WHO score, and a hypopharyngeal tumor site.

**Ethical Considerations:** The Regional Ethical Review Board in Gothenburg and Lund reviewed and approved the study, (Gothenburg, number 299–14, T230-17 and Lund 2020-01972). This study was carried out in accordance with the current Helsinki Declaration of the World Medical Association from 2013. Data are available upon request.

## Background

Head and neck cancer (HNC) is one of the most common cancers worldwide, and epidemiological assessments have reported a recent global increasing incidence[1–5], in Sweden as well as elsewhere over the recent years[6, 7]. Simultaneously, a reduced mortality has been noted in HNC in recent decades, due to better available treatments, less smoking, a multidisciplinary approach to treatment, and earlier discovery of tumors[8–11].

Numerous studies have dealt with survival in HNC patients, but population-based studies focusing on early death in HNC patients are scarce. Early death from HNC was previously defined as patients dying within six months of diagnosis in a study from the Swedish head and neck cancer register (SweHNCR)[12]. It reported that 4.5% of patients died within six months of diagnosis between 2008–2015, even though they received curative-intended treatment[12]. Curative-intended treatment is often aggressive, costly, and comes with considerable side effects. If patients receiving aggressive treatment die within six months, the physicians involved need to address whether the indications and decision-making involved in treatment management should be re-evaluated. Both surgery and radiotherapy, the backbones of head and neck cancer treatment, warrant repeated visits to the hospital and frequent hospitalization. Knowing the factors identifying patients at risk of early death could help clinicians better select suitable treatments, and thus avoid mutilating surgery and aggressive chemoradiotherapy in cases with a considerable risk of early death.

Since 2008, major progress in clinical management of patients with HNC has been made in Sweden. The care has, in general, been influenced by international initiatives aiming to improve quality of care of patients with HNC[5, 7]. Therefore, to study the efficacy of the new guidelines in an era of Human Papilloma Virus (HPV) and demographic changes, a further analysis of patients dying within six months of treatment is warranted. In this large, real-world, nationwide, population-based study containing 18,739 patients (16786 curative and 1769 palliative) with HNC, the primary aim was to identify patients at risk of early death after treatment with curative intent, and to analyze factors that contribute to this increased risk. Furthermore, we wanted to test the hypothesis as to whether six-month mortality has decreased for HNC patients over the past few years.

## Material and Methods

### Ethics statement

Data were obtained from the SweHNCR (Ethics Committee approval; Gothenburg, number 299–14, T230-17 and Lund 2020-01972). The SweHNCR is funded by the Swedish government and includes 98.5% of all HNC patients since 2008 when cross referenced with the register of the Department of Health and Welfare in Sweden. All data is anonymous. The authors did not have access to information that could identify individual participants during or after data collection. The total number of consecutively-affected Swedish patients during the period January 1, 2008–December 31, 2020 in the SweHNCR with at least six months of follow-up was 18,739[5].

### Patient population

Patients with a palliative treatment decision at multidisciplinary tumor board (MDT) (n =1953, 10.4%) were not included in the study, leaving 16,786 patients with curative treatment intent for further analysis. Patients with curatively intended treatment were divided into three groups based on year of diagnosis; 2008-2012, 2013-2016, and 2017-2020 (Figure 1). Ten sites of tumors were included: lip (C00.0–2, C00.6, C00.8, C00.9); oral cavity (C00.3, C00.4, C02, C03, C04, C05, C06); oropharynx (C01.9, C05.1, C05.2, C05.8, C05.9, C09, C10); nasopharynx (C11); hypopharynx (C12, C13); larynx (C10.1, C32); nose (C30.0) and nasal sinuses (C31); salivary glands (C07, C08); head and neck cancer of unknown primary (C77.0); and middle or inner ear (C 30.1). Malignant tumors located in the esophagus, thyroid, or parathyroid glands were not included. Data reported to the SweHNCR included: diagnosis, TNM classification (according TNM 7[13]), stage, sex, age, WHO score at diagnosis, time to treatment, treatment, follow-up, recurrence and survival. The Eastern Cooperative Oncology Group score, the WHO score, runs from 0 to 5, with 0 denoting perfect health and 5 death[14]. The WHO score was rated 0–4, indicating the physical performance status of the patient. A higher score indicates worse physical performance.

**Figure 1.**
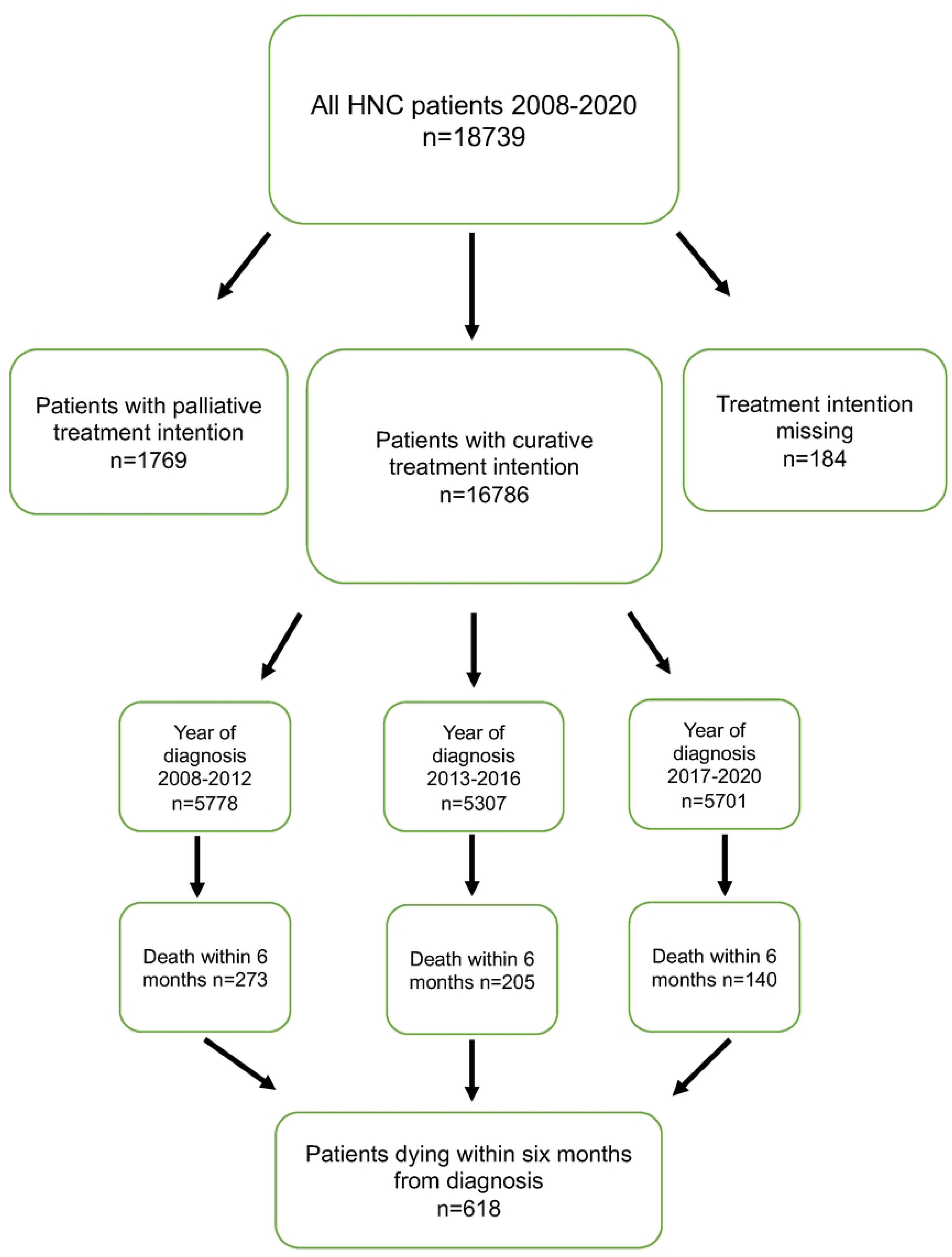
Flow chart describing treatment intent, year of diagnosis and early death for included patients.

HNC treatment in Sweden is centralized to seven university hospitals, although a few regional hospitals provide non-surgical oncological treatment. National Healthcare and Social Security systems are offered equally to all Swedish inhabitants. All patients are examined by either an oncologist or a head and neck surgeon after termination of treatment to evaluate treatment outcome, every three months for the first two years post treatment and every sixth months for the following three years.

### Statistics

Results are presented as the mean, standard deviation, and range for continuous variables, and as numbers and percentages for categorical variables. Incidence rates for 2008-2020 and incidence rate ratio was calculated comparing 2020 to 2008 with 95% confidence intervals according to a method described by Martin and Austin[6, 15–17]. Data from Statistics Sweden were used for information regarding yearly population count. SweHNCR was used to identify the number of new HNC cases annually. For comparisons between two groups (divided by; age, days to treatment, and days from diagnosis to death), the independent Student’s t-test was used. ANOVA test (with Bonferroni method as a post hoc test), the chi-square test for categorical variables (stage, site, age groups, time periods, and WHO function class) and Fisher’s exact test was used for tables with dichotomous variables (Sex, TNM score, and death). Exact binomial confidence intervals were estimated forproportions. Logistic regression analysis was used to control for confounding factors. To describe overall crude survival and early death for subgroups, a Kaplan-Meier plot was used, and the difference between subgroups (sex, time periods, age groups, and WHO class) was analyzed with a log-rank test. Cox regression was used for multivariable analyses with death within six months from diagnosis as failure. All significance tests were two-tailed and conducted at a 5% significance level. For all statistical analyses, we used IBM SPSS Statistics for Macintosh, Version 28.0. Armonk, NY: IBM Corp Released 2021.

## Results

The annual number of new patients with HNC steadily increased from 2008 (n=1211 cases) to 2020 (n=1673 cases). During the same period, the population increased by 12%, from 9.2M to 10.4M, indicating that factors other than a growing population were involved in the increase of new HNC cases. We noted an incidence rate ratio of 23.8 % (CI 95% 23.5-24.2) between 2008 (13.0 diagnosed with HNC per 100000) and 2020 (16.1 diagnosed with HNC per 100000), corresponding to an annual incidence increase of 0.25 per 100000 (p<0.001) (calculated as described in statistics section above) (Figure 2). Both oropharyngeal and oral cavity cancer displayed increased occurrence: the number of patients with oropharyngeal cancer rose with 86% from 2008 (n =270) to 2020 (n =503); oral cancer 48% from 2008 (n =343) to 2020 (n =509). The incidence rate ratio for oropharyngeal cancer was 66.1% (CI 95% 64.3-68.0) between 2008 (2.92 per 100000) and 2020 (4.85 per 100000), and for oral cavity cancer with 32.3% (CI 95% 32.0-33.6) from 2008 (3.71 per 100000) to 2020 (4.90 per 100000). All other sites had fairly similar occurrence over the years 2008-2020 (Figure 2). The number of patients discussed at multidisciplinary tumor boards (MDT) increased from 2008 to 2020 (95% to 99.5%, mean for the total study period 97%).

**Figure 2.**
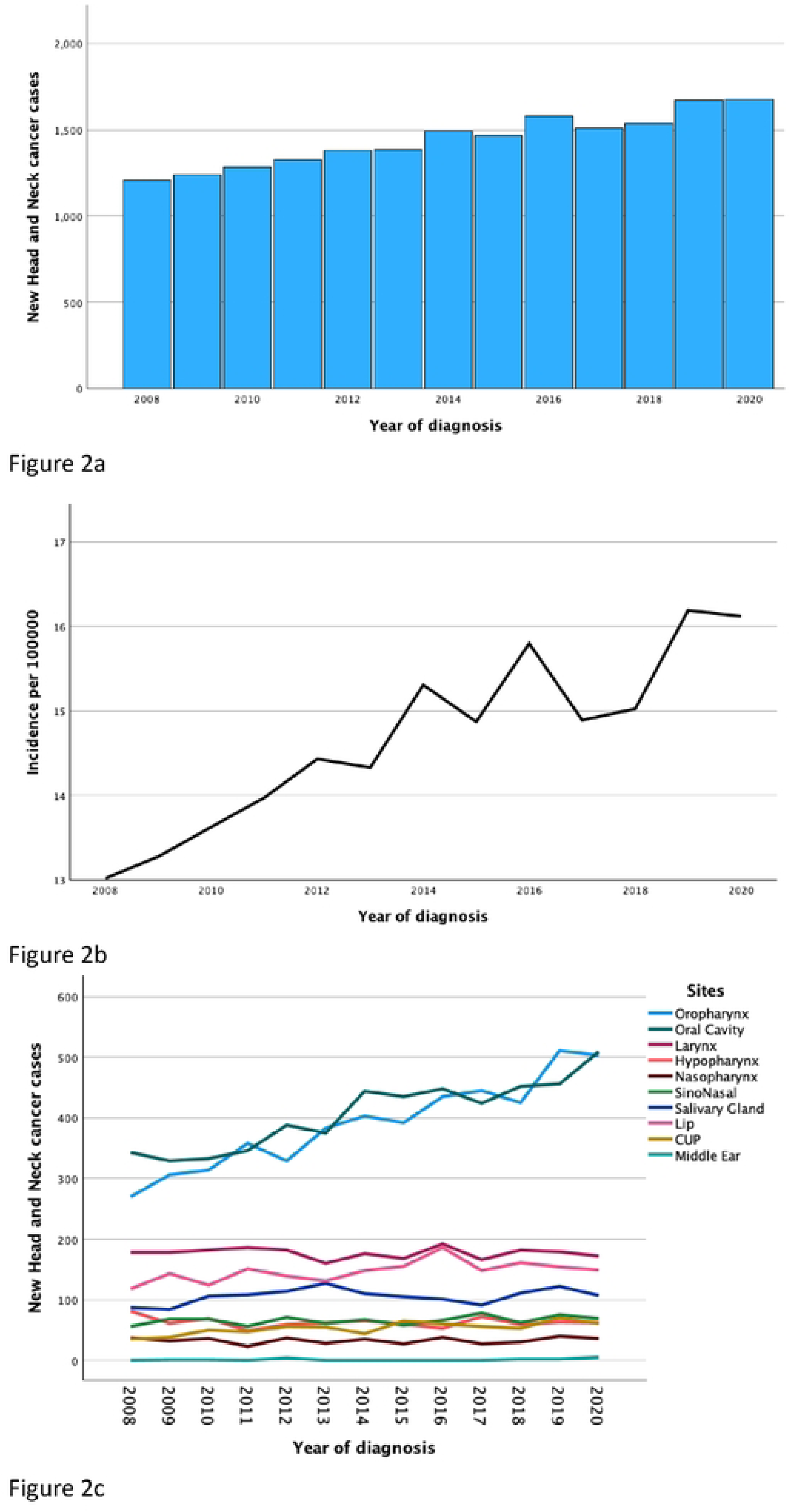
2a. Annual number of new cases of head and neck cancer in Sweden per year 2008-2020 2b. Incidence of head and neck cancer in Sweden 2008-2020 2c. Distribution of tumor sites of head and neck cancer in Sweden 2008-2020 Predictors for six-month mortality

During the years 2008 to 2020, 618 (3.7%) of 16,786 patients died within six months, despite having a recommendation for curatively intended treatment at MDT (Figure 1).

### Year of diagnosis

The total cohort was divided into three groups based on the year of diagnosis. The three groups contained a comparable number of patients: 2008–2012 (n=5778), 2013–2016 (n=5307), and 2017–2020 (n=5701) (Table 1, and 2). Between 2008–2012, six-month mortality was 4.7% compared to 3.9% and 2.5% for the years 2013–2016 and 2017–2020, respectively (p <0.001) (Table 1, Figure 3). Sex, age, and stage were evenly distributed throughout the three groups. The 2008–2012 group had a slightly higher distribution of unknown WHO scores compared to the two latter groups, p <0.001 (Table 2).

**Figure 3.**
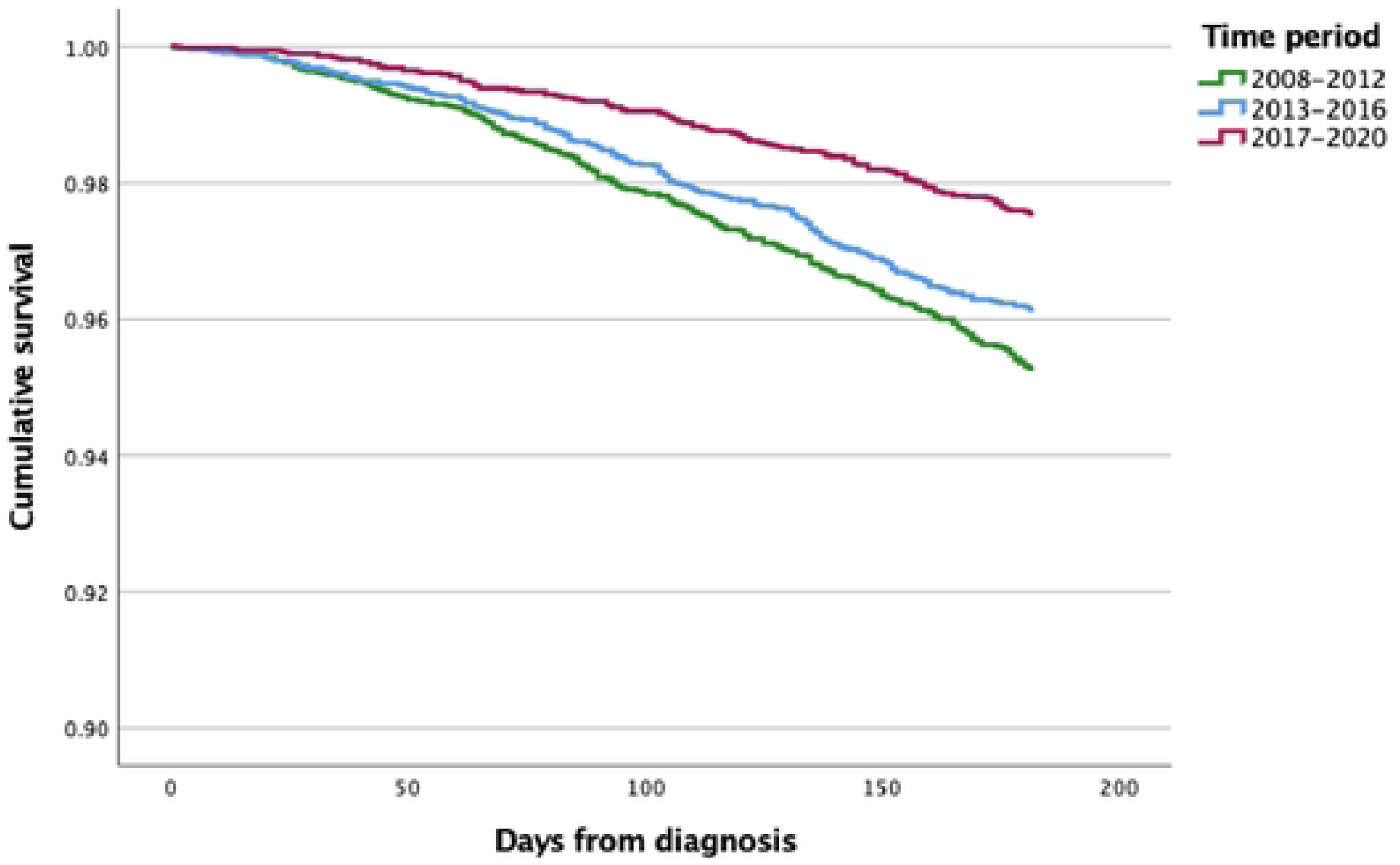
Kaplan-Meier curve of death within six months from diagnosis based on year of diagnosis

**Table 1.** Descriptive data for patients with curative treatment intent, and univariate analysis of death within 6 months.

| **N =16786** | **Prevalence n (%)** | **Dead within 6 months/N (%)** | **HR (95% CI)** | **P-values** |
| --- | --- | --- | --- | --- |
| **Sex** |  |  |  |  |
| Male | 10,712 (63.8) | 424/10,712 (4.0) | 1.00 | - |
| Female | 6074 (36.2) | 194/6074 (3.2) | 0.80 (0.68–0.95) | 0.012 |
| **Total** | 16,786(100) | 618/16,786(3.7) | - | - |
| **Age continuous (mean (sd))** | 66.1 (12.6) |  |  |  |
| **Age groups** |  |  |  |  |
| 18–39 | 463 (2.8) | 0/463 (0) | - | - |
| 40–49 | 1139 (6.8) | 14/1139 (1.2) | 1.00 | - |
| 50–59 | 3043 (18.1) | 52/3043 (1.7) | 1.40 (0.77–2.52) | 0.268 |
| 60–69 | 5378 (32.0) | 155/5378 (2.9) | 2.37 (1.37–4.10) | 0.002 |
| 70–79 | 4368 (26.0) | 203/4368 (4.6) | 3.86 (2.24–6.62) | <0.001 |
| 80+ | 2395 (14.3) | 194/2395 (8.1) | 6.88 (4.00–11.8) | <0.001 |
| **WHO Score** |  |  |  |  |
| 0 | 12,078 (72.0) | 206/12,078 (1.7) | 1.00 | - |
| 1 | 2244 (13.4) | 145/2244 (6.5) | 3.88 (3.14–4.80) | <0.001 |
| 2 | 890 (5.3) | 108/890 (12.1) | 7.53 (5.97–9.51) | <0.001 |
| 3 | 321 (1.9) | 68/321 (21.2) | 14.4 (11.0–19.0) | <0.001 |
| 4 | 36 (0.2) | 16/36 (44.4) | 37.2 (22.4–62.0) | <0.001 |
| Missing | 1217 (7.3) | 75/1217 (12.1) | 3.72 (2.85–4.84) | <0.001 |
| **Year of diagnosis** | 16,786 (100) | 618/16,786 (3.7) | - | - |
| 2008–2012 | 5778 (34.4) | 273/5778 (4.7) | 1.00 |  |
| 2013–2016 | 5307 (31.6) | 205/5307 (3.9) | 0.82 (0.68–0.98) | 0.026 |
| 2017–2020 | 5701 (34.0) | 140/5561 (2.5) | 0.51 (0.42–0.63) | <0.001 |

**Table 2.** Descriptive data for patients with curative treatment intent grouped by time of diagnosis and univariate analyses.

| Variable | 2008–2012 | 2013–2016 | 2017–2020 | Total<br>N (%) | Sig.<br>p-value |
| --- | --- | --- | --- | --- | --- |
| Oropharynx N (%) | 1461<br>(25.3) | 1499<br>(28.2) | 1738<br>(30.5) | 4698<br>(100) | <0.001 |
| Oral Cavity N (%) | 1531<br>(26.5) | 1514<br>(28.5) | 1615<br>(28.3) | 4660<br>(100) | <0.001 |
| Other sites N (%) | 2786<br>(48.2) | 2294<br>(43.2) | 2348<br>(41.2) | 7428<br>(100) | <0.001 |
| Male<br>N (%) | 3713<br>(64.3) | 3416<br>(64.4) | 3583<br>(62.8) | 10,712<br>(100) | 0.174 |
| Female<br>N (%) | 2065<br>(35.7) | 1891<br>(35.6) | 2118<br>(37.2) | 6074<br>(100) | 0.173 |
| WHO 0<br>N (%) | 4005<br>(69.3) | 3780<br>(71.2) | 4293<br>(75.3) | 12078<br>(72.0) | 0.013 |
| WHO 1<br>N (%) | 720<br>(12.5) | 763<br>(14.4) | 761<br>(13.3) | 224<br>(13.4) | 1.00* |
| WHO 2<br>N (%) | 289<br>(5.0) | 287<br>(5.4) | 314<br>(5.5) | 890<br>(5.3) | 0.544* |
| WHO 3<br>N (%) | 108<br>(1.9) | 95<br>(1.8) | 118<br>(2.1) | 321<br>(1.9) | 0.246* |
| WHO 4<br>N (%) | 14<br>(0.2) | 12<br>(0.2) | 10<br>(0.2) | 36<br>(0.2) | 0.604* |
| WHO missing<br>N (%) | 642<br>(11.1) | 370<br>(7.0) | 205<br>(3.6) | 1217<br>(7.3) | <0.001* |
| Total patients<br>N (%) | 5778<br>(100) | 5397<br>(100) | 5791<br>(100) | 16,786<br>(100) | - |
| Time to Surgery,<br>mean days*** | 22 | 19 | 14 | 19** | <0.001 |
| Time to Radiation<br>therapy, mean<br>days*** | 38 | 30 | 22 | 29** | <0.001 |

\*Chi-square between WHO1 and other WHO scores
\*\*Mean duration in days between 2008–2020
\*\*\*Days from diagnosis to start of treatment

The composition of oropharyngeal and oral cavity cancer differed between the 2017–2020 cohort (30.5% and 28.3% of all HNC cancer respectively) compared to the 2008–2012 cohort (25.3% and 26.3% respectively), p <0.001.

### Time to treatment

Time to the start of treatment from diagnosis was shorter in the 2017–2020 group compared to the group 2008–2012. The mean time to radiotherapy/chemoradiation therapy in the 2008–2012 cohort was 38 days, compared to 22 days for the 2017– 2020 cohort, p<0.001. The mean time to surgery from diagnosis was 22 days in 2008–2012, compared to 15 days for the 2017–2020 cohort, p <0.001 (Table 2).

### Sex

Altogether, 10,712 (63.8%) of patients were male, and 6074 (36.2%) were female (Table 1). A significant difference in mortality between the sexes was found. The six-month mortality rate among male patients was 4.0% compared to 3.2% for females (p =0.012) (Table 1) (Figure 4). Thus female patients had a 20% lower risk of death within six months compared to male patients.

**Figure 4.**
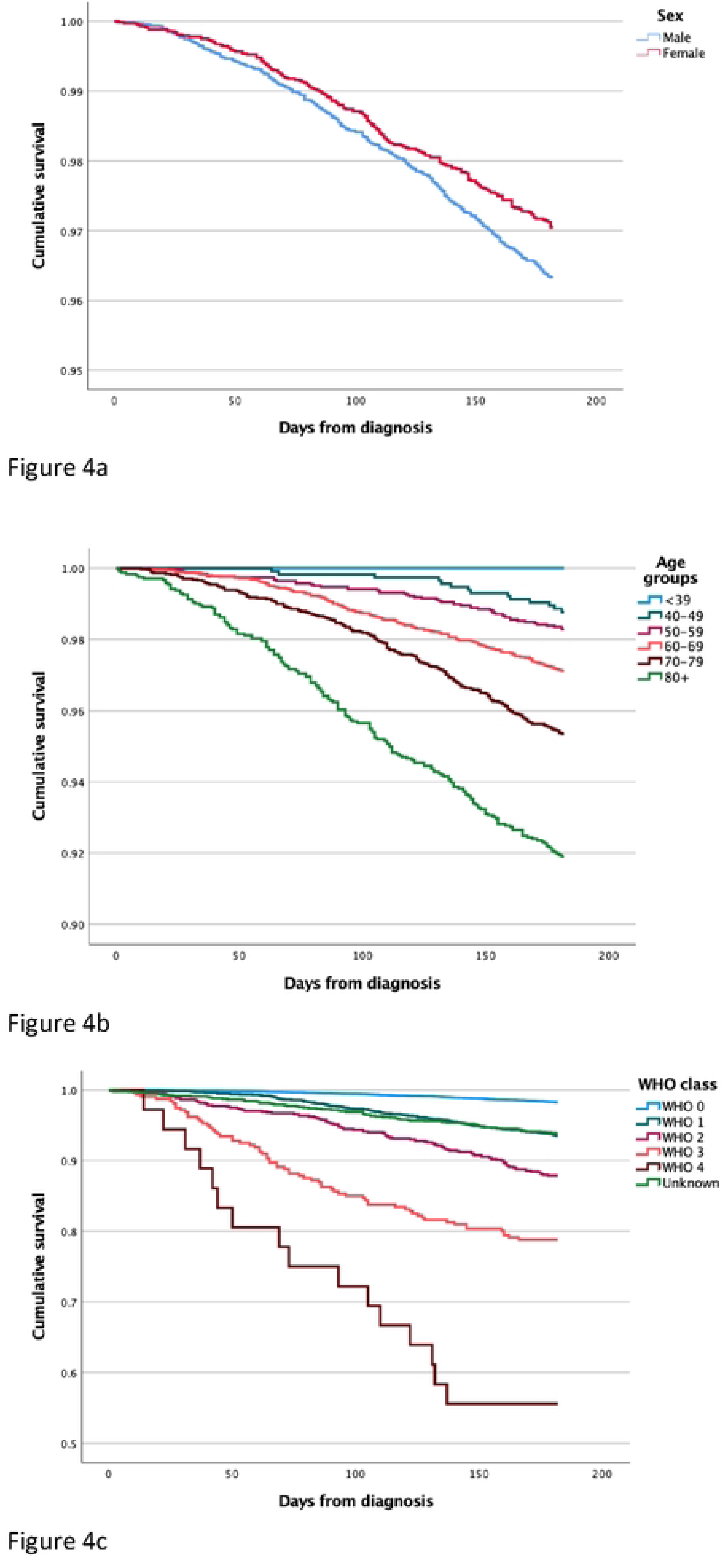
4a. Kaplan-Meier curve of death within six months from diagnosis based on sex 4b. Kaplan-Meier curve of death within six months from diagnosis based on age at diagnosis 4c. Kaplan-Meier curve of death within six months from diagnosis based on WHO function class at diagnosis

### Age

The mean age at diagnosis was 66 years (range: 18–102). Males were on average one year younger than the females (65.7 versus 66.7) at diagnosis. The risk of dying within six months increased with age. For every year older the patient was at diagnosis, six-month mortality increased by 4.8% (p <0.001). No patient below 40 years of age died within six months, compared to 8.1% of all patients older than 80 years (p <0.001). There was a 6.88-fold increased risk of dying within six months for patients over 80 years of age compared to patients below 50 years of age (Table 1) (Figure 4).

### WHO score

12,078 (72%) of all patients had a WHO score of 0. Only 1.7% of all patients with WHO 0 died within six months, compared to 44.4% of those with WHO 4 (p <0.001). In univariate analysis, the risk of death within six months was 37.2 times higher for patients with WHO 4 compared to those with WHO 0 (Table 1). Mean survival time among patients dying within six months was 119 days for WHO 0 patients compared to 76 days for WHO 4 patients (p =0.004) (Figure 4).

### Site

The most common tumor site was the oropharynx (28%) followed by the oral cavity (27.8%) and larynx (12.7%) (Table 3). The six-month mortality rate varied between tumor sites. Worst outcomes were seen in patients with hypopharyngeal cancer, with 10.3% dying within six months despite curative treatment intent. A univariable analysis showed that patients with a hypopharyngeal tumor had an almost 2.5-fold higher relative risk of dying within six months compared to oropharyngeal patients (p <0.001) (Table 3).

**Table 3.** Descriptive data for patients with curative treatment intent, and univariate analysis of death within 6 months.

| **N =16786** | **Prevalence n (%)** | **Dead within 6 months/N (%)** | **HR (95% CI)** | **P-value** |
| --- | --- | --- | --- | --- |
| **Tumor site** |  |  |  |  |
| Oropharynx | 4698 (28.0) | 146/4698 (3.1) | 1.00 | - |
| Oral Cavity | 4660 (27.8) | 201/4660 (4.3) | 1.40 (1.12–1.73) | 0.002 |
| Larynx | 2134 (12.7) | 89/2134 (4.2) | 1.35 (1.04–1.76) | 0.026 |
| Hypopharynx | 591 (3.5) | 61/591 (10.3) | 3.45 (2.56–4.65) | <0.001 |
| Nasopharynx | 381 (2.3) | 5/381 (1.3) | 0.42 (0.17–1.02) | 0.056 |
| Sino Nasal | 690 (4.1) | 26/690 (3.8) | 1.22 (0.80–1.85) | 0.355 |
| Salivary Gland | 1212 (7.2) | 35/1212 (2.9) | 0.93 (0.64–1.34) | 0.683 |
| Lip | 1850 (11.0) | 35/1850 (1.9) | 0.61 (0.42–0.88) | 0.008 |
| CUP | 559 (3.3) | 19/559 (3.4) | 1.10 (0.68–1.77) | 0.705 |
| Ear canal | 11 (0.1) | 1/10 (10) | 3.46 (0.48–24.7) | 0.216 |
| **Stage** |  |  |  |  |
| I | 4948 (29.5) | 73/4948 (1.5) | 1.00 | - |
| II | 2847 (17.0) | 66/2847 (2.3) | 1.58 (1.13–2.20) | 0.007 |
| III | 2077 (12.4) | 87/2077 (4.2) | 2.88 (2.11–3.93) | <0.001 |
| IV A | 5507 (32.8) | 293/5507 (5.3) | 3.68 (2.85–4.75) | <0.001 |
| IV B | 553 (3.3) | 56/553 (10.1) | 7.12 (5.03–10.1) | <0.001 |
| IV C | 93 (0.6) | 14/93 (15.1) | 11.0 (6.23–19.6) | <0.001 |
| Missing | 761 (4.5) | - | - | - |
| **T** |  |  |  |  |
| T1-2 | 11,649 (69.4) | 229/11,649 (2.0) | 1.00 | - |
| T3-4 | 4968 (29.6) | 383/4968 (7.7) | 4.04 (3.43–4.76) | <0.001 |
| Missing | 169 (1.0) | - | - | - |
| **N** |  |  |  |  |
| N- | 10,263 (61.2) | 300/10,263 (2.9) | 1.00 | - |
| N+ | 6520 (38.8) | 318/6520 (4.9) | 1.68 (1.44–1.97) | <0.001 |
| **M** |  |  |  |  |
| M0 | 16,511 (98.4) | 596/16,511 (3.6) | 1.00 | - |
| M1 | 102 (0.6) | 15/102 (14.7) | 4.36 (2.61–7.28) | <0.001 |
| Missing | 173 (1.0) | - | - | - |

### Stage

An association between tumor stage and six-month mortality was found. Among all patients in the total cohort, 54% were diagnosed with stage III or stage IV cancer (Table 3), although 55% of all women were diagnosed in stage I-II compared to 45% of all males (p<0.001). Of the 618 patients who died within 6 months, 76% had stage III or IV disease compared to 12% of patients with stage I. Significant differences in six-month mortality were also noted in stage IV. For stage I patients, six-month mortality was 1.5%, compared to 5.3%, 10.1%, and 15.1% for stages IV A, IV B, and IV C, respectively (p <0.001). In total, a patient with stage IV C disease had a 10-times higher risk of dying within six months compared to stage I patients (Table 3).

### TNM class

Higher T class correlated with worse prognosis. Seventy percent of all patients had a T1-2 tumor at diagnosis (Table 3). Only 2% of the patients with a T1-2 tumor died within six months compared to 7.7% of T3-4 patients. We found that a patient with T3-4 tumor had a 3 times higher risk of death within six months compared to T1-2 patients (p <0.001) (Table 3).

A total of 11,649 (61%) of all patients had no neck metastasis (N-) at diagnosis, whereas 6520 (39%) had one or more metastases in the neck at diagnosis (N+). The six-month mortality for N– patients was 2.9%, compared to 4.9% for N+ patients (p <0.001). In total, an N+ patient had a 1.68-fold higher risk of dying within six months compared to a N– patient (Table 3). One-hundred and two (0.6%) of all included patients had distant metastasis (M1) and were still considered at the MDT to benefit from curative treatment. However, 3.6% of patients with M0 disease died within six months, compared to 14.7% of patients with a M1 disease (p <0.001). We found that M1 patients had a 4.36-times higher risk of dying within six months compared to M0 patients (Table 3).

### Independent factors for death within six months after diagnosis

A multivariable Cox regression was carried out with death within six months as the dependent variable (Table 4). Patients diagnosed between the years 2008–2012 had a six-month mortality rate of 4.7% compared to 2.5% for patients diagnosed between 2017–2020, indicating a risk reduction of 53% (p <0.001) for death within six months (Table 4). Females had a 20% lower risk of dying within six months compared to males (p =0.013). For every year older the patient was at diagnosis, a 4.8% (p <0.001) higher risk of dying within six months was observed. A 67-year-old patient had a 4.8% higher risk of dying within six months than a patient who was 66 years old.

**Table 4.** Multivariable Cox regression with death within six months as target variable for HNC patients undergoing curative-intended treatment.

| Factor | HR (95% CI) | Significance |
| --- | --- | --- |
| **Year of diagnosis** |  |  |
| 2008–2012 | - | - |
| 2013–2016 | 0.83 (0.69–1.00) | 0.052 |
| 2017–2020 | 0.47 (0.40–0.61) | <0.001* |
| **TNM Score** |  |  |
| T1-2 vs 3-4 | 2.90 (2.43–3.51) | <0.001* |
| N+/0 | 1.60 (1.36–1.99) | <0.001* |
| M+/0 | 2.30 (1.36–3.85) | 0.002* |
| **Sex** | .800 (0.67–0.95) | 0.013* |
| **Age Continuously** | 1.05 (1.04–1.06) | <.001* |
| **WHO Function** |  |  |
| WHO 0 | - | - |
| WHO 1 | 2.38 (1.90–2.97) | <0.001* |
| WHO 2 | 3.84 (2.99–4.91) | <0.001* |
| WHO 3 | 7.87 (5.90–10.5) | <0.001* |
| WHO 4 | 27.6 (16.5–46.2) | <0.001* |
| WHO Missing | 2.90 (2.21–3.81) | <0.001* |
| **Tumor Site** |  |  |
| Oropharynx | - | - |
| Oral cavity | 1.21 (0.95–1.53) | 0.121 |
| Larynx | 1.13 (0.85–1.51) | 0.397 |
| Hypopharynx | 1.58 (1.16–2.14) | 0.003* |
| Nasopharynx | 0.510 (0.21–1.24) | 0.137 |
| Sino nasal | 0.984 (0.63–1.53) | 0.943 |
| Salivary Gland | 0.890 (0.61–1.31) | 0.555 |
| Lip | 0.629 (0.41–0.95) | 0.029* |
| CUP | 1.58 (0.95–2.64) | 0.077 |
| Ear | 4.84 (0.67–34.9) | 0.118 |

Patients with a WHO score of 1 had approximately 2.4-times greater risk of early death compared to WHO 0 patients (p <0.001). The risk of early death among WHO 4 patients was almost 28 times higher than for WHO 0 patients (p <0.001).

A patient with a tumor localized in the hypopharynx had a 1.58-times higher risk of dying within six months compared to a patient with a tumor in the oropharynx (p =0.003). Patients with a T3/4 tumor had a 2.9-times higher risk of dying within six months compared to T1/2 patients (p <0.001). It was found that patients with neck metastasis at diagnosis had a 1.6-times higher risk of dying within six months compared to patients without nodal involvement (p <0.001). Distant metastasis at diagnosis gave a 2.3-times higher risk of dying within six months (p =0.002) (Table 4).

These results form a risk profile where year of diagnosis, higher age, male sex, hypopharyngeal tumor, advanced TNM class, and higher WHO score constitute risk factors for early death.

## Discussion

This real-world, population-based, nationwide study, including 16,786 patients with HNC and curative treatment intent displayed that year of diagnosis, higher age, higher TNM class, male sex, higher WHO class, and a tumor in the hypopharynx were independent risk factors for death within six months of diagnosis. The effects of standardized protocols to ensure that all patients receive adequate treatment without unnecessary delays might have contributed to a significantly reduced risk of early death over recent years.

Throughout the study period, patients with curative treatment intent (n =16,786) had a 3.7% six-month mortality overall. Jensen et al. reported a 7.1% six-month mortality rate in a Danish nationwide study including 11,419 HNC patients between 2000–2017 treated with curatively intended radio- or chemoradiotherapy. The same author studied a series of 2209 HNC patients between 2010–2017 from multiple centers and found a non-cancer specific six-month mortality of 4.4% following treatment with radiotherapy[18, 19]. Nieminen et al. reported a six-month mortality of 11.4% in a subgroup of 317 HNC patients undergoing microvascular reconstruction[20]. As patients with HNC comprise a heterogenous population, the early mortality rate differs if subpopulations of patients are analyzed. Studies looking merely at treatment with radiotherapy, or at microvascular reconstruction contain selected patient cohorts likely affecting the rate of early death. Our study includes all surgical and oncological treatment options for patients with curative treatment intent and all T classes. Patients undergoing treatment with a free flap and microvascular reconstruction typically have at least a T2 tumor, indicating a disease with a higher risk of early death.

In the present study, men had a higher risk of early death, in accordance with the findings by Kouka et al. in their series of 8288 German HNC patients[21]. Other studies have not shown a sex-dependent rate of early death in HNC patients[12, 18]. In this study there is an uneven distribution between sexes regarding tumor stage and site; for example only 23.5% of hypopharyngeal cancer patients were female, which can partly explain the better prognostic outcome for females.

Other studies have suggested that estrogen could have a beneficial impact on survival after HNC [22–25], however that correlation was not analyzed here. In consistency with other studies [8, 12, 18, 20, 21, 26, 27], we saw that higher age and performance status correlated with an increased rate of early death. Furthermore, a hypopharyngeal tumor location, higher stage, and advanced T class were confirmed as significant independent risk factors for six-month mortality, in accordance with previous findings[12, 26, 28, 29].

Our study has revealed that the six-month mortality decreased throughout the period of 2008– 2020. The reasons are most likely multifactorial. Sweden implemented standardized diagnostic work ups in 2015, which led to faster diagnoses and shortened waiting times until treatment for HNC patients. The same year, national guidelines for HNC treatment were published, leading to a national standardized consensus regarding diagnostic work up and treatment[30]. Furthermore, oropharyngeal cancer derived from HPV is increasing [2, 7, 31–33], and has a better prognosis compared to HPV-negative tumors[34, 35]. The substantial increase in HPV-positive cancers and the decreased prevalence of daily smokers in Sweden (12% in 2008 to 8% in 2020), contributing to less HPV-negative HNC cases can be part of the explanation as to why six-month mortality is decreasing for HNC patients [36–38].

It is reasonable to speculate that a number of variables not registered in the SweHNCR have improved the diagnostic work-up, treatment, and care of patients with HNC. Neck dissection was implemented 2015 in Sweden as the standard treatment for oral cavity cancer patients with a depth of invasion of the primary tumor exceeding 3 millimeters and N0 status. This change is likely contributing to more active treatment of occult metastasis, and therefore to improved survival rates. It is also likely that new innovative therapy has had an impact on early mortality. Since 2008, there have been several improvements in oncological treatments such as the change of external radiotherapy from using 3D conformal radiotherapy to intensity-modulated radiotherapy and volumetric modulated arc radiotherapy, shifts from neoadjuvant high-dose cisplatin regimens to concomitant cisplatin. Concomitant cetuximab treatment was proven to be less effective, and was excluded from routine treatment[39]. New diagnostic tools have been increasingly available for accurate HNC diagnosis. The implementation of sentinel node biopsies in 2018 could also have contributed to identifying more contra- or bilateral metastases. Moreover, the use of FDG-PET/CT has improved the detection of regional and distant metastases. Altogether, this study suggests that standardized and rapid diagnostic work up and treatment, together with other factors, contributes to fewer HNC patients dying within six months of diagnosis.

Weaknesses of the study includes the fact that important parameters influencing overall survival, such as comorbidities, alcohol consumption, HPV status, and smoking habits were not recorded in the early years of the SweHNCR. Actual cause of death was not reported in the register, along with data on occupational exposures, socioeconomic factors, oral hygiene, complications to treatments, and gene mutations; all factors which could influence early mortality[1, 27, 40, 41].

Compensating for several of the study’s limitations is the large population of 16,786 consecutive HNC patients with curative treatment intent, and the homogeneity of healthcare provided to all Swedes. The results in this study can be used to identify HNC patients at high risk of early death and thus open up for discussions about de-escalation of treatment for selected cases to provide the best possible remaining time for patients at high risk of early death.

## Conclusion

This first European population-based analysis of trends in six-month mortality among HNC patients with curative treatment intent is based on registry data comprising 16,786 patients. The study provides generalizable epidemiological data on early mortality and survival of curative HNC patients. The mean time to treatment was significantly decreased between the years 2008-2012 compared to 2017-2020. Encouragingly, the present results show that early mortality has decreased from 4.7% (2008–2012) to 2.5% (2017–2020) in Sweden. Female patients had a 20% lower risk of death within six months compared to male patients. Increased age at diagnosis, male sex, higher WHO function class, advanced TNM class, and tumor in the hypopharynx were independent risk factors for death within six months.

## Data Availability

All relevant data are within the manuscript and its Supporting Information files.

## Acknowledgements

We would like to thank Mats Fredriksson, statistician at Linköping University, for excellent statistical guidance.

